# Clinical description, molecular delineation and genotype-phenotype correlation in 340 patients with KBG syndrome: Addition of 67 new patients

**DOI:** 10.1101/2022.04.11.22271283

**Authors:** Elena Martínez-Cayuelas, Fiona Blanco-Kelly, Fermina López-Grondona, Saoud Tahsin-Swafiri, Rosario López-Rodríguez, Rebeca Losada-Del Pozo, Ignacio Mahillo, Beatriz Moreno, María Rodrigo-Moreno, Dídac Casas-Alba, Aitor López-González, Sixto García-Miñaur, María de los Ángeles Mori, Marta Pacio-Mínguez, Emi Rikeros-Orozco, Fernando Santos-Simarro, Jaime Cruz-Rojo, Juan Francisco Quesada-Espinosa, María Teresa Sánchez-Calvin, Jaime Sánchez-del Pozo, Raquel Bernadó-Fonz, María Isidoro-García, Irene Ruiz-Ayucar, María Isabel Álvarez, Raquel Blanco-Lago, Begoña De Azua-Brea, Jesús Eirís, Juan José García-Peñas, Belén Gil- Fournier, Carmen Gómez-Lado, Nadia Irazabal, Vanessa López, Irene Madrigal, Ignacio Málaga, Beatriz Martínez-Menéndez, María Soraya Ramiro-Leon, María García-Hoyos, Pablo Prieto-Matos, Javier López-Pisón, Sergio Aguilera-Albesa, Sara Álvarez de Andrés, Alberto Fernández-Jaén, Isabel Llano-Rivas, Blanca Gener, Carmen Ayuso, Ana Arteche-López, María Palomares-Bralo, Anna Cueto, Irene Valenzuela, Antonio F. Martínez-Monseny, Isabel Lorda-Sánchez, Berta Almoguera

**Affiliations:** Department of Pediatrics. Fundación Jiménez Díaz University Hospital; Department of Genetics and Genomics, IIS–Fundación Jiménez Díaz University Hospital; Center for Biomedical Network Research on Rare Diseases (CIBERER), ISCIII, Madrid, Spain; Department of Statistics. IIS-Fundación Jiménez Díaz University Hospital; Clinical Genetics and Dysmorphology. Department of Genetic and Molecular Medicine. Pediatric Institute of Rare Diseases (IPER) Sant Joan de Deu Hospital, Barcelona; Medical and Molecular Genetics Institute (INGEMM), La Paz University Hospital, IdiPaz; ITHACA- European Reference Network, Spain; UDISGEN (Dysmorphology and Genetics Unit), 12 de Octubre University Hospital, Madrid; Department of Pediatrics, Endocrinology Unit, 12 de Octubre University Hospital, Madrid, Spain; Department of Genetics. 12 de Octubre University Hospital, Madrid, Spain; Pediatric Neurology Unit. Department of Pediatrics. Navarra University Hospital. Navarrabiomed Pediatric Neurology Research Group; Department of Biochemistry. Salamanca University Hospital; Department of Pediatrics. Salamanca University Hospital; Department of Biochemistry and Molecular Genetics, Clinic Hospital, IDIBAPS (Institut de Investigacions Biomèdiques August Pi I Sunyer); Pediatric Neurology Unit. Department of Pediatrics. Central de Asturias University Hospital; Department of Pediatrics. Son Llátzer University Hospital; Department of Pediatric Neurology. Santiago University Hospital; Pediatric Neurology Unit. Department of Pediatrics. Niño Jesús University Hospital; Department of Genetics. Getafe University Hospital; Department of Pediatrics. Can Misses University Hospital; Medical Genetics Unit. Department of Genetics. Virgen de la Arrixaca University Hospital. IMIB-Arrixaca; Pediatric Neurology Unit. Department of Neurology. Getafe University Hospital; Genomics and Medicine, NIMGenetics. Madrid; Department of Pediatrics. Miguel Servet University Hospital; Department of Pediatric Neurology. Hospital Universitario Quirónsalud. Madrid. Spain; School of Medicine. European University of Madrid; Department of Genetics. Cruces University Hospital; Department of Clinical and Molecular Genetics, Vall d Hebron University Hospital and Medicine Genetics Group, Vall d Hebron Research Institute, Barcelona, Spain

## Abstract

**Background:** KBG syndrome is a highly variable neurodevelopmental disorder and clinical diagnostic criteria have changed as new patients have been published. Both loss-of-function sequence variants and large deletions (CNVs) involving *ANKRD11* have been involved in KBG, but no genotype-phenotype correlation has been reported to date. This study presents the clinical and molecular characteristics of 67 new patients with KBG syndrome and the results of the first genotype-phenotype correlation leveraging data on 273 patients previously published.

**Methods:** 67 patients with KBG syndrome were recruited through a Spanish collaborative effort and were assessed using a custom phenotypic questionnaire. The frequency of all features was calculated. Manifestations present in >50% of the patients and a “severity score” were used to perform a genotype-phenotype correlation in the 340 KBG patients.

**Results:** Neurodevelopmental delay (95%), comorbidites (82.8%), macrodontia (80.9%), triangular face (71%), characteristic ears (76%), nose (75.9%) and eyebrows (67.3%) were the most prevalent features in the 67 patients. The genotype-phenotype correlation yielded significant associations with the triangular face (71.1% in patients with sequence variants vs 45.2% in CNVs, p=0.015), short stature (62.5% variants in exon 9 vs. 27.8% outside; p=0.009) and macrodontia (with larger deletions, p=0.028), ID/ADHD/ASD (70.4% in c.1903_1907del vs. 89.4%; p=0.012) and a higher phenotypic score in patients with sequence variants compared with CNVs (p=0.005).

**Conclusions:** We present a detailed phenotypic description of KBG syndrome in the largest series of patients reported to date, provide evidence of a genotype-phenotype correlation between some KBG features and specific *ANKRD11* aberrations, and propose updated clinical diagnostic criteria based on our findings.

## INTRODUCTION

KBG syndrome (MIM: 148050) is caused by haploinsufficiency of Ankyrin Repeat Domain 11 (*ANKRD11)*, an important chromatin regulator, capable of inducing changes in gene expression. *ANKRD11* plays an important role in neurodevelopment as it acts as a regulator in the developing brain, where it influences proliferation of neural progenitors, the genesis and positioning of newborn neurons, neuronal plasticity and dendritic differentiation, playing an important role in neurodevelopment[1–3].

In the last decade, *ANKRD11* has been associated with intellectual disability (ID), facial dysmorphism and developmental disorders, with more than 300 patients with KBG syndrome reported in the literature to date (Human Genetic Mutation Database HGMD professional version; accessed in February 2022). Indeed, the incidence of KBG syndrome is suggested to be underestimated due to the variability in the severity and manifestations of the phenotype[4, 5]

The cardinal features of the syndrome are developmental delay (DD), ID, and a characteristic skeletal and facial phenotype consistent of postnatal short stature, hand and costovertebral anomalies, macrodontia of the upper central incisors and a triangular face[6–9]. Additional manifestations such as hearing loss and seizures are frequent comorbidities in KBG patients. However, the clinical presentation of the syndrome is highly heterogeneous regarding both the phenotype and severity of the symptoms, with variable expressivity and penetrance[10]. This has resulted in a lack of a consensus clinical diagnostic criteria, with manifestations considered major and minor changing as new patients have been published[6–9, 11–13].

Skjei and colleagues were the first to propose a diagnostic criteria of KBG based on a series of 46 patients[11]. According to their proposal, patients had to have DD/ID and four out of eight major findings: macrodontia, a characteristic facial appearance, hand anomalies, costovertebral anomalies, delayed bone age (<2SD), short stature (<p3), neurological involvement and first degree relative with KBG[11]. With the publication of additional series of patients, this initial criteria evolved to one based on four major (macrodontia or characteristic facial gestalt, postnatal stature below the 10th centile, recurrent otitis media/hearing loss, and a first degree relative with KBG) and seven minor (hand anomalies, seizures, cryptorchidism, feeding problems, palate abnormalities, a formal diagnosis of autism spectrum disorder -ASD- and a wide/large fontanelle)[8]. Based on these criteria, patients had to have at least two major or one major plus two minor features in order to meet a clinical diagnosis of KBG syndrome[8].

Both *de novo* and dominantly inherited loss-of-function variants in *ANKRD11*, primarily in exon 9, and large deletions in 16q24.3 have been identified in KBG patients (HGMD professional version, accessed in February 2020). Several authors have attempted to establish a genotype-phenotype correlation with the position or type of *ANKRD11* aberration[7, 8, 14]. Although preliminary evidence has suggested associations with neurological involvement, congenital heart defects, vision problems or severe otitis media, no statistical significance has been reached, possibly due to the limited number of patients studied[7, 8, 14]. In this study, we present the clinical and molecular data of 67 new patients with KBG syndrome, which is the largest series of patients described to date, along with 273 patients previously reported in the literature. Data from these 340 patients were used to delineate the clinical characteristics of KBG syndrome and their frequency, and to perform the first genotype-phenotype correlation.

## MATERIALS AND METHODS

### Selection of KBG patients and collection of phenotypic information

The project was approved by the ethics committee of all participating sites and was performed in accordance with the Declaration of Helsinki Principles and institutional requirements. Written informed consent for genetic tests, for data sharing and publication of identifiable patient images was obtained from each participant or their guardians.

Patients were recruited through a national collaborative effort involving 19 centers of expertise in pediatric neurology, rare diseases, and clinical genetics in Spain (Supplementary Material). The selection of subjects was performed by retrospective review of patients with a diagnosis of KBG syndrome due to pathogenic sequence variants or large deletions involving *ANKRD11*.

All KBG patients were examined by a clinical geneticist and/or a pediatric neurologist, who collected phenotypic information from each participant. The phenotype information was collected using a custom deep phenotyping questionnaire developed for this project (Supplementary Material) based on existing phenotypic information of KBG syndrome in the literature and additional clinical features deemed of interest by the collaborators. Categories included in the questionnaire were: 1. Demographic data; 2. Personal history; 3. Neurodevelopment; 4. Neurological findings; 5. Other clinical findings; 6. Physical examination; 7. Medical investigations; and 8. Genetic investigation and findings (Supplementary Material).

### Genetic tests and variant analysis

Tests performed for the genetic diagnosis of KBG included the following: 1. Clinical exome sequencing using the Clinical Exome Solution v2 of 4900 genes by Sophia Genetics (CES; Sophia Genetics, Boston, MA) and Illumina True Sight One with 6713 genes (TSO; Illumina, San Diego, CA). 2. Array CGH using the aCGX 60K platform (CGXTM, PerkinElmer, Inc), CytoScan. 3. Direct Sanger sequencing of *ANKRD11*. 4. Whole exome sequencing, with the Agilent SureSelect v6 library, the Agilent SureSelect Clinical Research Exome (Agilent Santa Clara, CA), or the Ion AmpliSeq(tm) Exome Kit (Life Technologies). 5. Whole genome sequencing. Variant interpretation and classification followed the American College of Medical Genetics and Genomics guidelines for *ANKRD11* sequence variants and the International Standards for Cytogenomic Arrays for large deletions. All variants were referred to the genomic construct hg19 and *ANKRD11* transcript NM_013275.6.

### Literature review

A PubMed search was conducted to select papers reporting patients with KBG syndrome and pathogenic variants in *ANKRD11* from 2010 to December 2021. Inclusion criteria was based on fulfillment of the search terms “KBG syndrome” OR “*ANKRD11*” and only papers in English, reporting pathogenic variants, and with a comprehensive description of the phenotype were considered. Patients with missense *ANKRD11* variants not previously reported as pathogenic or not confirmed as *de novo* were discarded. These patients were used to 1) describe the spectrum of pathogenic variants and their prevalence, 2) describe the clinical features of the syndrome, their frequency, and compare with data from our cohort; and 3) to perform genotype-phenotype correlations.

For all patients selected by the literature review, phenotypic and molecular information included in the questionnaire was extracted and converted to the cutoffs established. Presence and absence of the features were considered when there was a specific mention of that presence or absence or when patients were documented to be assessed for that specific feature. Not mentioning a given feature was considered as a missing value except in large series with frequencies calculated from the totals, where patients with no mention of the feature were considered negative for that feature.

### Data analysis

#### Variables

Phenotypic information from the questionnaire was reviewed and variables with missingness ≤60% in our cohort of patients were excluded or grouped. Frequency of each feature in the two cohorts separately and combined was calculated. Age of patients, age for first words, and age at independent walking was expressed as mean (± standard deviation, SD). Language and motor delays were considered when having first words and when independent walking was achieved after 18 months of age, respectively. ID was graded into four categories: borderline, mild, moderate, and severe. ASD and Attention deficit hyperactivity disorder (ADHD) categories included patients with either a formal diagnosis or features of such conditions.

Growth measurements of length, weight, and head circumference in our cohort were expressed as percentiles. Postnatal microcephaly and short stature were defined as measurements <p3 and <p10, respectively. Data from the literature were converted to the same cutoffs.

#### Statistical analyses

##### Comparison of KBG syndrome features between our cohort and patients from the literature

Frequency of phenotypic features from the questionnaire in our cohort and from the literature was calculated and compared using a Chi squared test or Fisher’s exact test. Significance was set as p<0.05. Analyses were performed using the R software v4.0.5.

##### Genotype-phenotype correlation

Phenotypic features present in more than 50% of the patients were selected for the genotype-phenotype correlation and grouped as follows: 1. History of developmental delay (motor and/or language delay); 2. ID/ASD/ADHD (ID, ASD, and/or ADHD); 3. Main comorbidities: seizures, congenital heart defects, hearing loss, recurrent otitis media, visual problems, feeding difficulties, and cryptorchidism. 4. Phenotypic features: macrodontia and other dental anomalies; triangular face; long philtrum; characteristic nose (anteverted nares, bulbous tip, and/or prominent nose), eyebrows (wide and/or thick and/or bushy and/or synophrys), ears (large and/or prominent and/or low-set); hand anomalies (brachydactyly and/or clinodactyly); and postnatal short stature (<p10). For binary variables (presence or absence of the feature), presence was defined as having at least one of the features defining the group, and for the comorbidities the total number per patient was calculated. A “phenotypic score” was created for the genotype-phenotype correlation, as a measure of the severity and complexity of the syndrome.

The severity score consisted of the sum of the 17 features from the four groups described above: history of developmental delay, ID/ASD/ADHD, presence of seizures, congenital heart defects, hearing loss, recurrent otitis media, visual problems, feeding difficulties, and cryptorchidism; macrodontia and other dental anomalies; triangular face; long philtrum; characteristic nose, eyebrows, and ears; hand anomalies; and postnatal short stature. Presence of each of the variables scored 1, so the maximum value of the phenotypic score was 17. Patients with more than 60% of missing values were excluded from the analysis.

Genetic variants in *ANKRD11* were classified into large deletions (CNVs) and sequence *ANKRD11* variants, and the latter were further subdivided according to whether they fell inside or outside exon 9, or whether it was the recurrent c.1903_1907del variant.

For the genotype-phenotype correlation, each individual feature, the number of comorbidities, and the phenotypic score were compared in the following groups: patients with sequence variants vs CNVs, variants inside and outside exon 9, c.1903_1907del vs other sequence variants and deletion size in CNV carriers. For the correlation with binary variables, proportions were compared with a Chi squared or Fisher’s exact tests. For the correlation with the continuous variables (number of comorbidities and phenotypic score), median values and quartiles were compared with a Mann-Whitney U test. For the comparison of the deletion size with the quantitative variables, a Spearman correlation coefficient was used. Statistical significance was set as p<0.05.

## RESULTS

### Patients and variables included in the study

Sixty seven patients from 64 families were recruited from 19 different centers across Spain (Supplementary Material): 39 patients were males and 28 were females, with a mean age (±SD) of 14.7 (±8.1) years (range of 2-50 years) and 84.1% were Caucasians. The suspected clinical diagnosis was primarily KBG (N=18), followed by Cornelia de Lange (N=11) and Kabuki syndromes (N=4) (some patients had more than one suspected diagnosis).

The literature review yielded 295 patients from 42 papers (see Supplementary Material). Six of the 295 patients were excluded due to having a missense variant in *ANKRD11* not confirmed *de novo*, and 16 patients lacked comprehensive phenotypic data, and were also excluded. The final number of patients included was 273 from 40 studies (Supplementary Material)[6, 7, 18–27, 8, 28–37, 9, 38–46, 12–17]

All 340 patients (67 from our cohort and 273 from the literature) were evaluated using the questionnaire developed for this study and data were converted according to the cutoffs established (Supplementary Material).

### Demographic and clinical description of 67 new patients with KBG syndrome and comparison with 273 patients from the literature

A summary of the clinical and phenotypic data from our cohort is shown in Table 1, along with data from the 273 previously published KBG patients and the frequency from all patients combined. Extended data for the 340 patients can be found in the Supplementary Material (Tables S1 and S2)

**Table 1.**
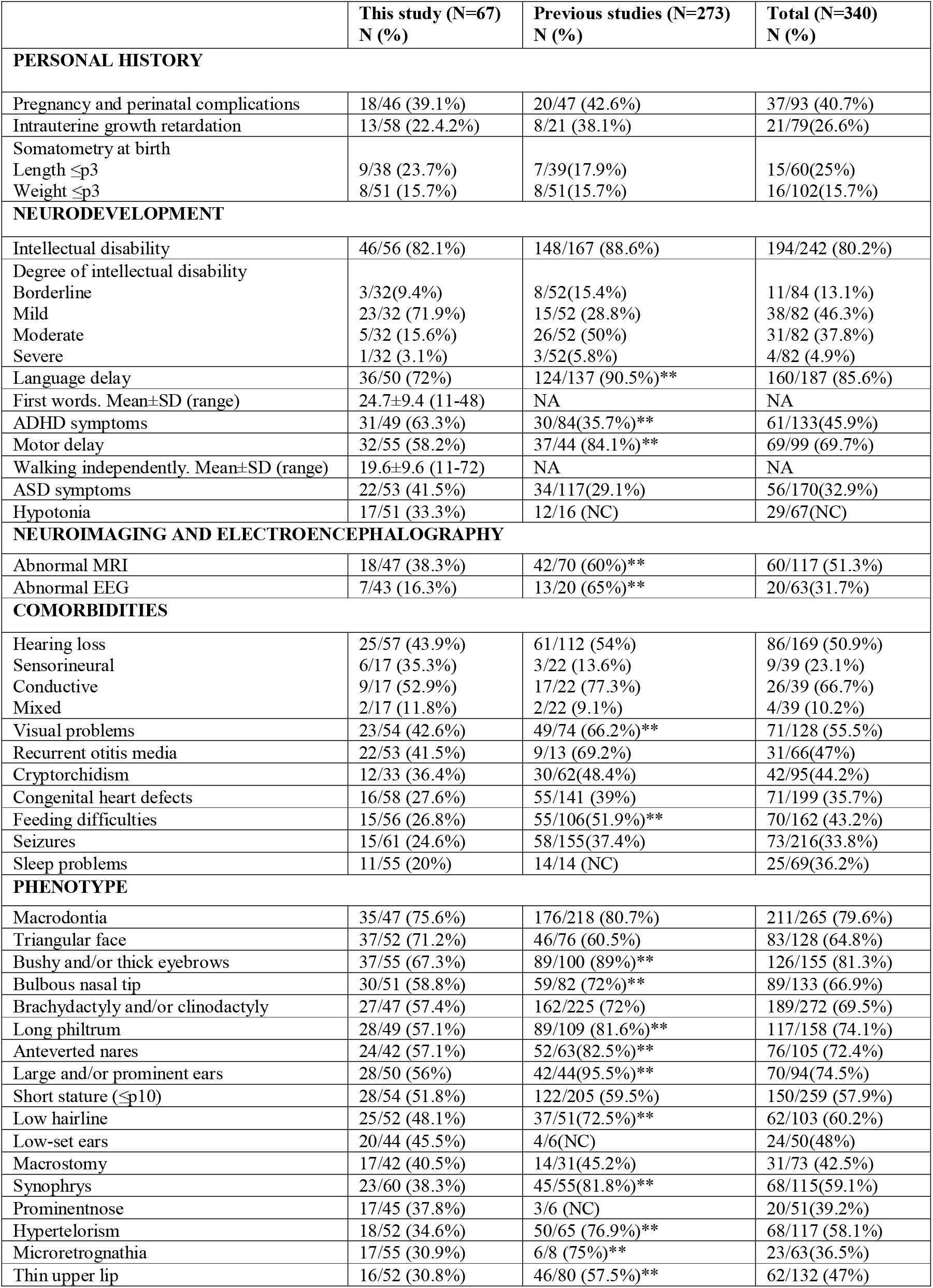
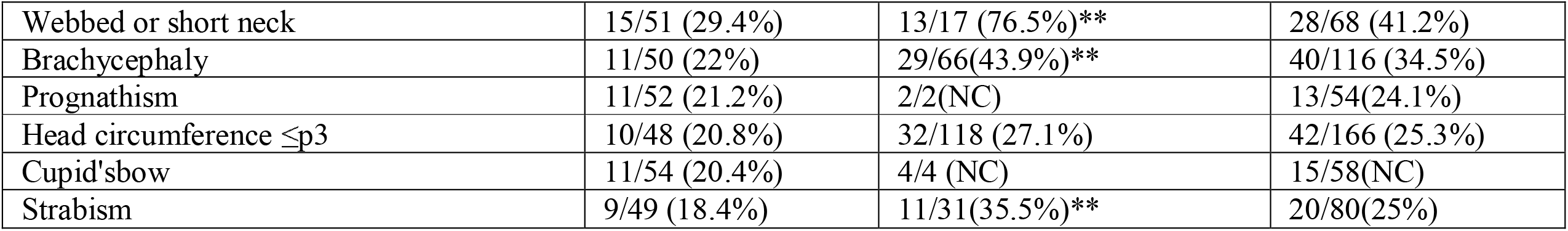
Clinical characteristics of KBG patients from our series, the literature and all combined. Age at walking and first words, expressed in months. Frequency of features was determined considering only patients assessed for each specific feature, indicated in the denominator. NA=not available; NC=not calculated. **refers to a statistically significant difference between the two series of patients (p<0.05).

Pregnancy and perinatal complications were present in 39.1% of the patients, with intrauterine growth retardation in 22.4% and weight and length at birth ≤p3 in 23.7% and 15.7% of patients, respectively.

Fifty eight percent of patients had motor delay, 72% language delay, and 88% had ID (82.1%), and/or ADHD (63.3%), or ASD (41.5%) symptoms. When the main comorbidities were considered together, 82.8% of patients had either hearing loss and/or otitis media; cardiopathy (27.6%; most commonly ventricular septal defect), seizures (24.6%: six focal, four generalized, four febrile seizures and two electrical status epilepticus in sleep); visual problems (42.6%; primarily refractive errors -81.8%- and strabismus -18.4%-). Cryptorchidism was reported in 36.4% of the male patients and two unrelated patients had a double excretory system. Regarding the phenotype, the most common features were macrodontia (75.6%), a characteristic nose (bulbous nasal tip, anteverted nares and/or prominent nose, 78.9%); a triangular face (71.2%); thick eyebrows and/or synophrys (67.3%); characteristic ears (large and/or prominent and/or low-set, 64%), hand anomalies (brachydactyly and/or clinodactyly, 57.4%); and a long philtrum (57.1%). Brain MRI was abnormal in 38.3% of patients, who had unspecific findings: enlargement of cisterna magna (9%), leukoencephalopathy (4.5%), anomalous cerebellum (4.5%), ventriculomegaly (3%), and anomalies of corpus callosum (3%), and/or hyppoccampus (1.5%).

The frequency of some of the features described (N=18) was significantly lower (p<0.05) in our cohort of patients compared to those from the literature: language delay (78% vs 90.5%); visual problems (40.7% vs 66.2%); feeding difficulties (26.8% vs 51.9%); MRI (38.3% vs 60%) and EEG (16.3% vs 65%) abnormalities; and 15 of the 24 phenotypic features (Table 1). The frequency of ADHD symptoms was significantly higher in our cohort of patients (63.3% vs 35.7%).

### 3. Molecular findings on the 67 new patients and 273 patients from the literature

Fifty eight of the 67 patients from our cohort (86.6%) carried 44 different heterozygous pathogenic sequence variants in *ANKRD11* (Table 2) and nine (13.8%) carried a deletion at 16q24.3 involving *ANKRD11* (Table 3). Of the 44 pathogenic sequence variants, 33 were novel (75%) and eleven had been previously reported (Table 2), and were mostly located in exon 9 (79.5%). There were six recurrent sequence variants: c.1903_1907del; p.Lys635GInfs*26 found in seven unrelated patients, c.6792dup; p.Ala2265Argfs*8 found in four patients from three families; and c.2329_2332del; p.Glu777Argfs*5; c.4384dup; p.Arg1462Lysfs*92; and c.5790C>A; p.Tyr1930* found in two unrelated patients each. Information about the origin of the variant was available for 39 of the 67 patients and was *de novo* in 36 cases (92.3%) and inherited from an affected mother in three patients (KBG8A, KBG10A, and KBG31A).

**Table 2.**
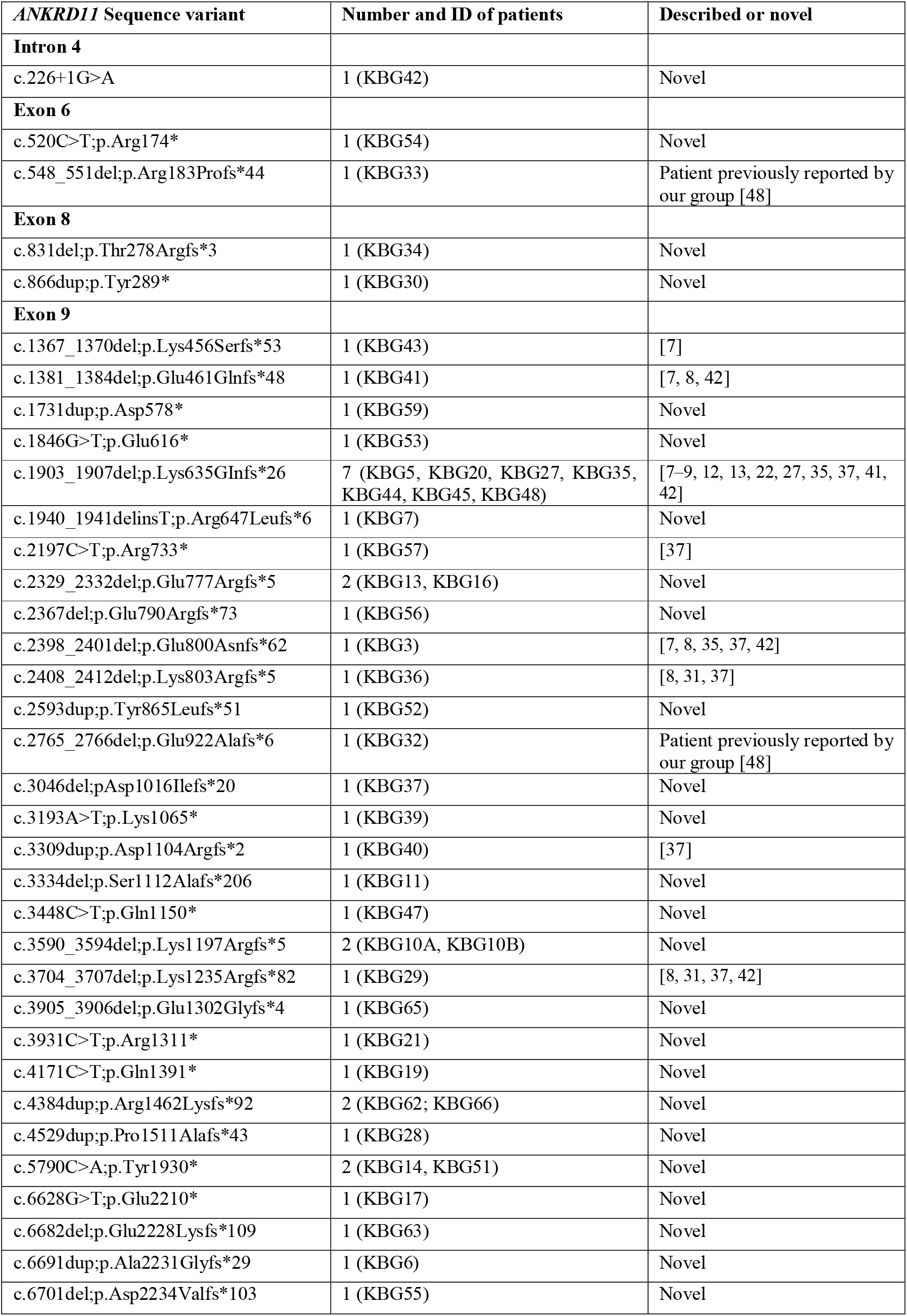
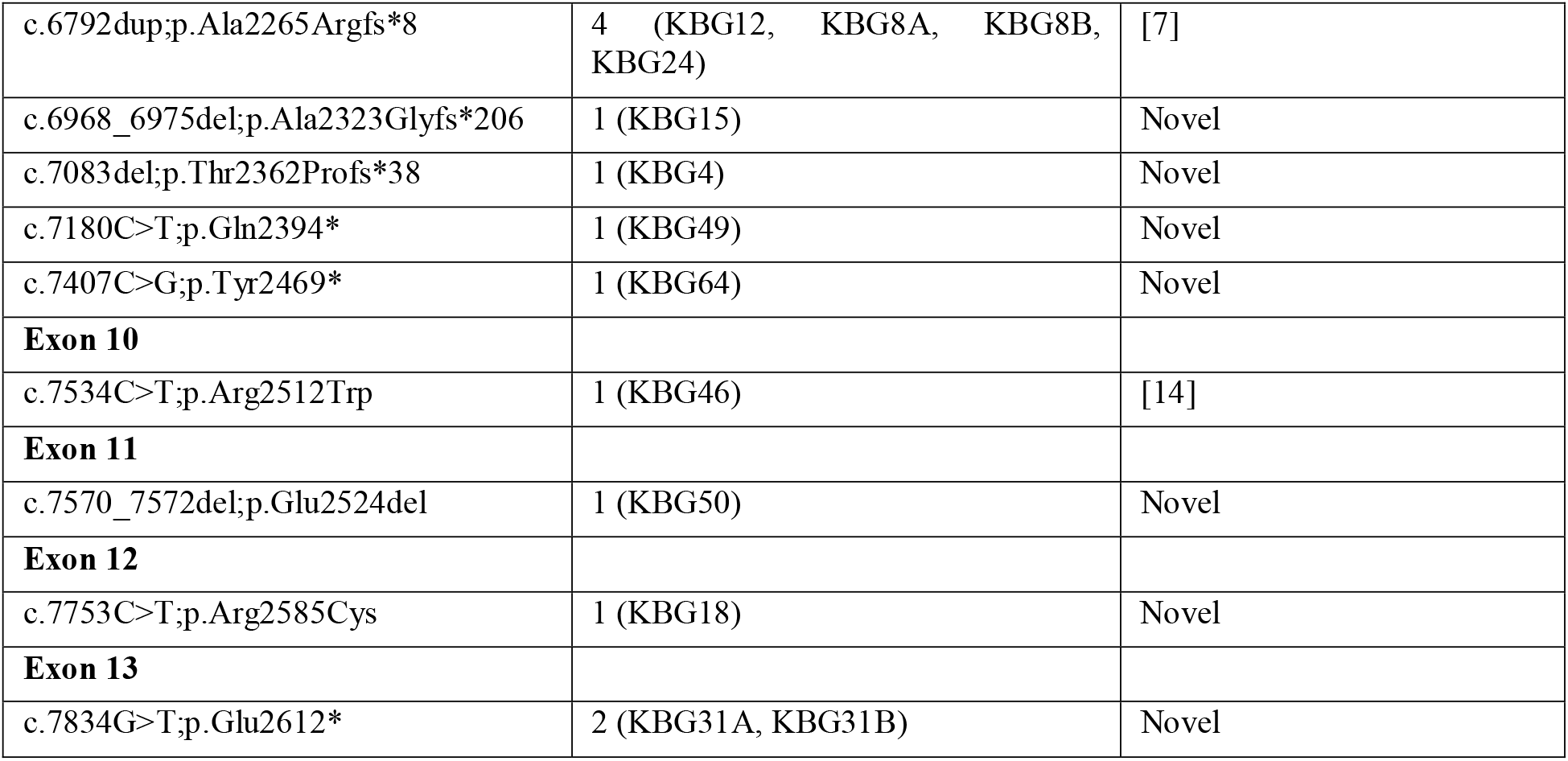
Pathogenic *ANKRD11* sequence variants identified in 58 patients sorted by exon, the number and identifier of patients (ID) and whether the variant is novel or not. *ANKRD11* transcript used is NM_013275.

**Table 3.**
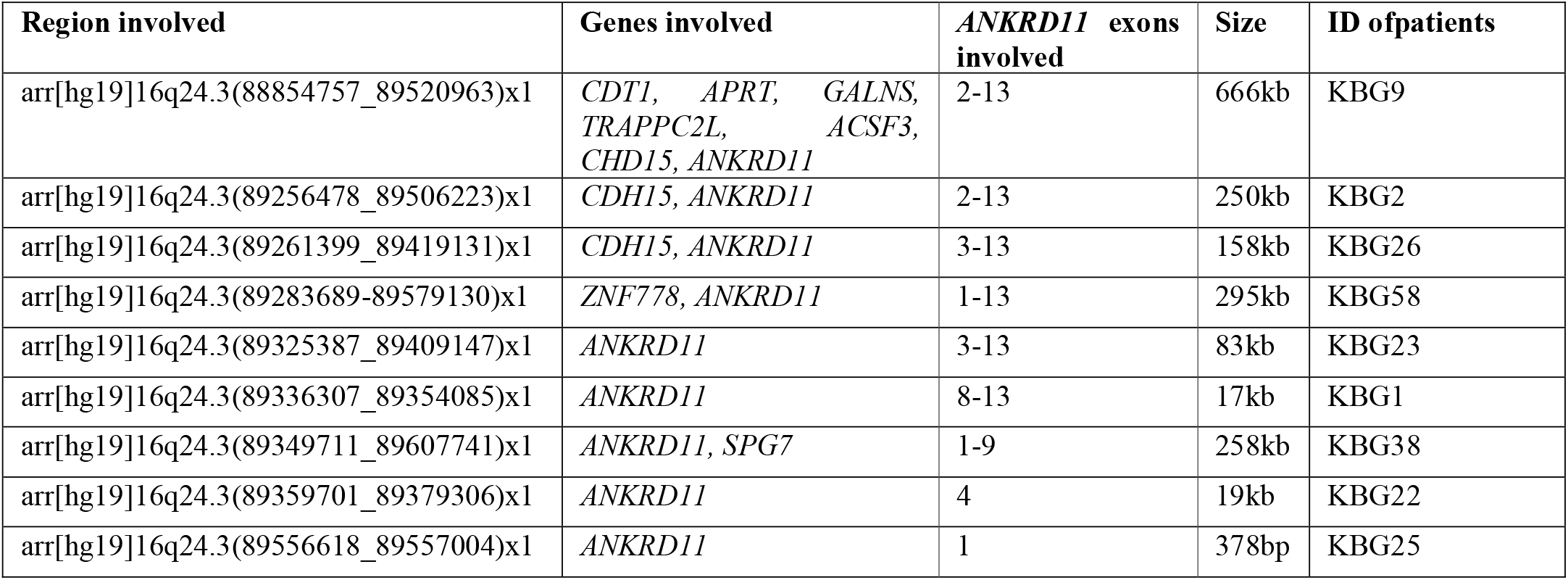
16q24.3 deletions identified in nine patients with the genes included in the deleted region, the *ANKRD11* exons affected, the size of the variant and the ID of the patient. Genomic coordinates refer to hg19 and *ANKRD11* transcript used is NM_013275.

Overall, of the entire cohort of 340 patients with KBG syndrome, 78.5% had a pathogenic sequence variant in *ANKRD11*, and four were recurrent: c.1903_1907del; p.Lys635GInfs*26 identified in 34 patients (10% of all KBG patients), c.2408_2412del; p.Lys803Argfs*5 in 10 (2.9%); c.1381_1384del; p.Glu461Glnfs*48 in 8 (2.4%); and c.2398_2401del; p.Glu800Asnfs*62 in 6 patients (1.8%), which explained 17% of all cases with KBG syndrome.

### 4. Genotype-phenotype correlation

For the genotype-phenotype correlation, the frequency of the features with a frequency >50% in the patients (Table 4) and the phenotypic score were compared in carriers of SNVs vs CNVs; carriers of variants inside and outside exon 9, carriers and non-carriers of the c.1903_1907del and correlated with the deletion size, in CNV carriers.

**Table 4.** Categories of variables with a frequency>50% in our series of KBG patients defining the phenotypic score used for the genotype-phenotype correlation

| Category | Variable | Our series<br>N= 67 patients | Literature<br>N= 273 patients | All<br>N= 340 patients |
| --- | --- | --- | --- | --- |
| Neurodevelopment | Motor and/or language delay | 86.5% | 99.2% | 95.7% |
|  | ID and/or ADHD and/or ASD | 95% | 85.8% | 88.0% |
| Phenotype | Characteristic nose | 75.9% | 96.8% | 91.2% |
|  | Macrodonia and/or other dental anomalies | 80.9% | 84.6% | 83.9% |
|  | Characteristic eyebrows | 67.3% | 89.0% | 81.3% |
|  | Long philtrum | 51.8% | 95.8% | 79.6% |
|  | Characteristic ears | 76.0% | 83.0% | 79.4% |
|  | Hand anomalies (brachydactyly and/or clinodactyly) | 57.5% | 71.2% | 69.5% |
|  | Triangular face | 71.2% | 60.5% | 64.8% |
|  | Short stature | 51.9% | 59.5% | 57.9% |
| Comorbidities | Any of the seven main comorbidities | 82.8% | 94.2% | 91.5% |
|  | Hearing loss and /or otitis media | 55.6% | 95.7% | 78.2% |
|  | Other comorbidities | 75.0% | 91.5% | 87.4% |

The phenotypic score was calculated with 17 variables: ten from the neurodevelopment and phenotype categories and the seven main comorbidities (Supplementary Material Table S3). The median value of the score was 6 ranging from 0 to 15.

The genotype-phenotype correlation yielded various significant associations (Table 5). Triangular face was significantly higher in patients with sequence variants compared to CNVs (71.1% vs. 45.2%, respectively; p=0.015) and short stature was significantly more frequent in patients with variants in exon 9 than outside (62.5% vs. 27.8%, respectively, p=0.009). Patients carrying the c.1903_1907del variant had lower frequency of ID/ADHD/ASD (70.4% vs. 89.4%, respectively; p=0.012). The size of the deletion was significantly associated with the presence of macrodontia and/or dental anomalies, more frequent in larger deletions (p=0.028); and with hand anomalies, with the opposite effect (p=0.048). Finally, the phenotypic score was significantly higher in patients with sequence variants compared with CNVs (median of 7 vs 6, p=0.005)

**Table 5.** Phenotype-genotype correlation of the most frequent features of KBG syndrome and the phenotypic score. Four groups of comparisons are shown: sequence variants versus CNV; sequence variants inside and outside exon 9; recurrent c.1903_1907del variant versus other sequence variants; and deletion size in carriers of CNVs. For CNV deletion size, Yes and No refers to patients having and not having the specific feature. For the first three groups, binary variables are expressed as frequency and continuous as median and quartiles (1^st^, 3^rd^ quartile) of the number of comorbidities or score. For the CNV deletion size, binary variables are expressed as median and quartiles (1^st^, 3^rd^ quartile) of the CNV size and quantitative variables are expressed with the Spearmann correlation score.

| Variable | Sequence variants | CNV | P-value | Exon 9 | No exon 9 | P-value | c.1903_1907del | Other variant | p-value | Deletion size |  | p-value |
| --- | --- | --- | --- | --- | --- | --- | --- | --- | --- | --- | --- | --- |
|  |  |  |  |  |  |  |  |  |  | Yes | No |  |
| Developmental delay | 134 (96.4%) | 42 (93.2%) | 0.407 | 124 (96.1%) | 10 (100%) | 1.000 | 14 (93.3%) | 120 (96.8%) | 0.440 | 240 (144, 795) | 295 (272, 480) | 0.625 |
| ID/ADHD/ASD | 180 (99.4%) | 40 (93.0%) | 0.392 | 159 (85.5%) | 21 (100%) | 0.083 | 19 (70.4%) | 161 (89.4%) | 0.012* | 250 (142, 479) | 666 (333, 1283) | 0.720 |
| Triangular face | 69 (71.1%) | 14 (45.2%) | 0.015* | 60 (71.4%) | 9 (69.2%) | 1.000 | 6 (75.0%) | 63 (70.8%) | 1.000 | 250 (202, 334) | 690 (158, 1000) | 0.074 |
| Macrodonia/dental anomalies | 182 (84.7%) | 42 (80.8%) | 0.636 | 165 (84.6%) | 17 (85.0%) | 1.000 | 23 (85.2%) | 159 (84.6%) | 1.000 | 256 (149, 609) | 97 (39, 154) | 0.028* |
| Long philtrum | 103 (79.8%) | 18 (78.3%) | 0.786 | 93 (79.5%) | 10 (83.3%) | 1.000 | 11 (73.3%) | 92 (80.7%) | 0.502 | 154 (94, 582) | 250 (158, 295) | 0.890 |
| Characteristic nose | 113 (91.1%) | 21 (91.3%) | 1.000 | 102 (91.1%) | 11 (91.7%) | 1.000 | 13 (81.2%) | 100 (92.6%) | 0.152 | 249 (157, 434) | 333 (167, 500) | 0.641 |
| Characteristic eyebrows | 105 (82.0%) | 21 (77.8%) | 0.808 | 98 (83.1%) | 7 (70.0%) | 0.384 | 10 (71.4%) | 95 (83.3%) | 0.278 | 274 (142, 603) | 748 (285, 957) | 0.280 |
| Characteristic ears | 59 (83.1%) | 18 (69.2%) | 0.225 | 49 (81.7%) | 10 (90.9%) | 0.676 | 6 (75.0%) | 53 (84.1%) | 0.615 | 268 (140, 371) | 466 (233, 1025) | 0.267 |
| Hand anomalies | 157 (72.4%) | 32 (58.2%) | 0.061 | 140 (70.7%) | 17 (89.5%) | 0.139 | 22 (78.6%) | 135 (71.4%) | 0.574 | 223 (128, 357) | 580 (156, 1100) | 0.048 |
| Short stature | 115 (59.3%) | 35 (53.8%) | 0.534 | 110 (62.5%) | 5 (27.8%) | 0.009* | 14 (53.8%) | 101 (60.1%) | 0.696 | 232 (135, 732) | 264 (128, 854) | 0.818 |
| Hearing loss/otitis media | 75 (77.3%) | 22 (81.5%) | 0.842 | 65 (77.4%) | 10 (76.9%) | 1.000 | 7 (70.0%) | 68 (78.2%) | 0.690 | 240 (149, 952) | 224 (116, 388) | 0.522 |
| Other comorbidities | 174 (86.1%) | 47 (92.2%) | 0.358 | 157 (85.8%) | 17 (89.5%) | 1.000 | 21 (87.5%) | 153 (86.0%) | 1.000 | 332 (152, 1025) | 83 (42, 118) | 0.039 |
| Number of comorbidities | 1.0 (1.0, 2.0) | 1.0 (1.0, 2.0) | 0.585 | 1.0 (1.0, 2.0) | 2.0 (1.0, 2.2) | 1.000 | 1.0 (1.0, 2.0) | 1.0 (1.0, 2.0) | 0.582 | 0.09 |  | 0.542 |
| Phenotypic score | 7.0 (6.0, 8.0) | 6.0 (4.0, 8.0) | 0.005* | 7.0 (6.0, 8.0) | 7.0 (5.8, 9.2) | 0.461 | 7.0 (6.0, 8.2) | 7.0 (5.5, 8.0) | 0.665 | -0.27 |  | 0.059 |

## DISCUSSION

In this study, we present the clinical and molecular characteristics of the largest series of 67 patients with KBG syndrome described to date. Using a comprehensive phenotypic questionnaire in our cohort and in 273 KBG patients previously published in the literature, we provide a detailed description of the syndrome and the results of the first genotype-phenotype correlation.

In our cohort, ID was the most common neurodevelopmental finding (82.1%), along with language delay (72%) and ADHD diagnosis or symptoms (63.3%), especially milder forms, which is in line with previous reports [5, 12–14, 45, 49]. We included patients who met all clinical criteria for ASD and ADHD, and therefore had a formal diagnosis, but also those with clear symptoms related to those entities, despite not meeting all criteria for a definitive diagnosis. We decided to be more inclusive in the definition of ASD and ADHD to better describe the spectrum of symptoms in KBG syndrome. This could explain the high prevalence of ADHD in our sample compared to that from the 273 patients from the literature. Nevertheless, a recent study by Kutkowska-Kazmierczak and colleagues described that 86% of the 23 patients reported had “psychomotor hyperactivity” and proposed to include these symptoms in the diagnostic criteria of KBG syndrome[13].

On the other hand, 41.5% of our patients had ASD diagnosis or symptoms, which is similar to the frequency reported by Ockeloen and colleagues[12], but higher than that found in other large series of patients[7–9, 14, 37], and that the frequency we determined from patients from the literature (29.1%).

Regarding the phenotype, there was a large interindividual variability in our patients but macrodontia, triangular face, bushy eyebrows and a long philtrum were features consistently found. Other common features were brachydactyly and/or clinodactyly, anteverted nares, a bulbous nasal tip, and large or prominent ears, all characteristic of the syndrome[6–8, 12]. Notably, 53 of 64 patients (83%) had comorbidities. The most common was hearing loss and/or otitis media, present in 55.6% of the patients, followed by visual problems, cryptorchidism, congenital heart defects, feeding difficulties, or seizures. Even though epilepsy and EEG anomalies have been frequently reported in previous series of KBG syndrome, no specific epileptic profile has been described[10, 12, 14, 37, 44]. In our study, both focal and generalized seizures were present; two individuals presented electrical status epilepticus in sleep which, to our knowledge, has not been previously reported, and four had febrile seizures, which has just been recently described[44].

When the frequency of KBG features was compared between our sample and patients from the literature, there were some variables that were significantly lower in our series, such as motor or language delay; visual problems, feeding difficulties, abnormal MRI or EEG, as well as several phenotypic features. Differences in the definition or age cutoffs for language or motor delay; a non-active search of some co-morbid complications such as feeding difficulties, visual problems or MRI/EEG abnormalities; the difficulty in the detection of dysmorphic facial features, which may be subtle and age-dependent; and a bias in the reporting of some features could explain such differences. Indeed, most variables displaying differences between the two cohorts were reported in a small proportion of the patients in the literature and we only considered absence of the feature when the report specifically stated that absence or the entire cohort had been assessed for the feature. This criterion, along with the systematic phenotypic assessment in our cohort of patients by the use of a questionnaire, may result in an artificially higher frequency of such variables compared to that seen in our sample.

The phenotypic evaluation of patients was performed in our sample using a specific and comprehensive phenotypic questionnaire, which allowed for a systematic assessment of features in our cohort. We believe that the frequency of KBG features yielded from our cohort is more accurate due to the large sample size and the systematic assessment of each specific feature, which allows a more objective evaluation of the patients. We encourage the use of this questionnaire or a similar tool for the phenotypic evaluation of KBG patients as it would aid in the delineation and recognition of the syndrome and ultimately in the diagnosis of the patients.

Both large deletions and sequence variants in *ANKRD11* cause KBG syndrome[10]. Despite the attempt of several authors to establish a correlation between the phenotype of the patients and the position or type of *ANKRD11* aberration, no significant associations have been reported to date [7–9, 12]. This has likely been the consequence of the limited sample sizes investigated[7–9, 12]. To overcome such limitation, we combined genetic and phenotypic data from our 67 patients with that of 273 patients previously published in the literature with a comprehensive phenotype description. We explored associations with phenotypic features found with a frequency over 50% in the entire cohort of 340 patients, and aggregated into categories, and with a phenotypic score, which was created as a measure of the complexity and severity of the phenotype. Such variables were compared between patients with sequence variants vs CNVs, sequence variants inside and outside exon 9, patients with the recurrent c.1903_1907del variant and in carriers of CNVs, associated with the deletion size. There was a significantly higher frequency of patients with a triangular face in carriers of sequence variants compared to CNVs, who also had significantly higher phenotypic score, suggesting the occurrence of a more severe or syndromic phenotype in patients with sequence variants compared to CNVs. Other associations found were short stature and variants in exon 9, a lower incidence of ID/ADHD/ASD in carriers of the c.1903_1907del variant and the size of the deletion, in CNV carriers, with the presence of macrodontia and hand anomalies. Although these findings may need to be further confirmed, the results of the genotype-phenotype correlation support the existence of distinctive findings in different *ANKRD11* variant types.

As previously stated, and evidenced by our data, there is wide phenotypic heterogeneity in KBG syndrome regarding the manifestations and severity of the phenotype[10]. This heterogeneity has resulted in the update of the diagnostic criteria of the syndrome as new patients have been published[7, 8, 11, 12]. The first diagnostic criteria, proposed by Skjei, included having DD and/or ID and four of the following eight findings: macrodontia, a characteristic facial appearance, hand anomalies, costovertebral anomalies, delayed bone age, short stature, neurological involvement and a first degree relative with KBG[11]. The studies led by Goldenberg and Ockeloen proposed to remove delayed bone age and costovertebral anomalies from the criteria and to not consider macrodontia mandatory[7, 12]. The latest diagnostic criteria by Low and colleagues in 2016, however, considered macrodontia as one major criteria and they also included recurrent otitis media and/or hearing loss as major criteria, adding the presence of seizures, cryptorchidism, feeding problems, palate abnormalities, a formal diagnosis of ASD and a wide/large fontanelle as minor criteria, not including delayed bone age and costovertebral anomalies[8].

Data from our series of patients supports the removal of delayed bone age and costovertebral anomalies as they are not systematically investigated (assessed in 28 and 39 of 67 patients, respectively) although, in our opinion, after diagnosis, costovertebral anomalies should be ruled out to discard possible complications. Also, we think that macrodontia must be kept as major since, besides being highly prevalent in KBG patients (80.9% in our series), is one of the most characteristic features of the syndrome, often clue to diagnosis and easy to explore. Furthermore, although some of the facial features of KBG patients could be subtle, others may be easily recognizable and useful in the differential diagnosis of the syndrome. Thus, we propose to use facial features as a criterion independent of macrodontia, especially those related to ears (76%), nose (75.9%), and eyebrows (67.3%), as well as the triangular face (71.2%), as they are highly prevalent among patients. In addition, macrodontia of upper central incisors could not be present before the eruption of permanent teeth so that, in children younger than six years old, facial phenotype, developmental delay and comorbidities, would help in establishing the diagnosis[37].

Hearing loss and/or otitis media, considered by Low as a major criterion[8] was present in 55.6% of the 67 patients. However, when any of the main seven comorbidities was considered, 82.8% of the patients had hearing loss and/or otitis media, congenital heart defects, seizures, vision problems, feeding difficulties, and/or cryptorchidism. Thus, we strongly suggest considering the presence of any of such main comorbidities as major criterion in the diagnosis of KBG.

Having a first-degree relative with a diagnosis of KBG syndrome was also proposed as major criterion by Low and colleagues[8]. Despite this fact being probably the most suggestive finding of KBG syndrome, it would only be met in a small proportion of patients, as pathogenic variants in *ANKRD11* are *de novo* in most patients (86% of the 340 cases).

ASD proposed by Low as part of the diagnostic criteria[8] was substantially lower than ADHD in our series (41.5% vs 63.6%). Indeed, of the 58 patients with information on neurodevelopment (ADHD/ID/ASD), 57 had either ID or ADHD with or without ASD (98.3%), and only one had a diagnosis of ASD as the main neurodevelopmental condition (Supplementary Material). Therefore, we recommend using ADHD symptomatology as a criterion instead of or in addition to ASD.

Lastly, fontanelle and palatal abnormalities have been found with variable frequencies in KBG syndrome, but as high as 22% and 58% of the patients, respectively[8]. In our series, only 4 of 31 patients (12.9%) had a wide anterior fontanelle with delayed closure compared to the 38% calculated from the literature (29 of 76 patients from the total of 273) (data not shown), and none were found to have palatal abnormalities. Those differences can be explained by the fact that these features were not systematically explored in the patients. More evidence is needed to determine their real frequency in KBG patients.

The diagnostic criteria proposed by Low in our cohort were met by 70% of our KBG syndrome patients (data not shown). Therefore, we calculated the percentage of patients who would meet the diagnostic criteria for KBG considering the categories illustrated in Table 4: 88.7% of the patients had DD and/or ID/ADHD/ASD and either ≥3 phenotypic features or ≥1 phenotypic features plus ≥1 main comorbidity (Supplementary Material, Table S4). Therefore, we propose having either a history of neurodevelopmental delay (motor and/or language) or ID, ADHD and/or ASD and at least three phenotypic features or less than three plus, at least, one of the seven main comorbidities as diagnostic criteria for KBG.

This study has some limitations that are intrinsic to the retrospective review of patients, both from our cohort and the literature, and the criteria used for data management, which leads to a considerable proportion of missing data that may impact the results. However, compared to previous works, this study has strengths that, in our opinion, outweigh the limitations. First, the sample size described, which is the largest to date, and the comprehensive phenotypic assessment made with a custom questionnaire that was used in our 67 patients and the 273 from the literature. The exhaustive literature review performed in this work and the extraction of the genetic and comprehensive phenotypic data on 340 patients using the same questionnaire have made it possible to provide a detailed phenotypic description and frequencies of the main features of KBG syndrome, and to perform the first genotype-phenotype correlation.

## Data Availability

All data produced in the present work are contained in the manuscript

## ACKNOWLEDGMENTS

The authors would like to thank all patients and their parents, as well as the Spanish Association of KBG syndrome, for their participation in the study. We thank Fernando Infantes, Inés García, Ascensión Giménez, Miguel Ángel López, Jesús Gallego, Camilo Vélez, Rocío Cardero, Laura Horcajada, and the Raregenomics Network (S2017/BMD-3721) for their technical and intellectual support.

## ETHICS

The study protocol was approved by the institutional review board (EO165-19_FJD, approval date 22/10/2019).

## CONTRIBUTORSHIP STATEMENT

E.M-C, F.B-K, I.L.S. and B.A. designed the study. E.M-C, F.B-K, R.L-R, I.M, I.L.S, and B.A. analyzed the data; and E.M-C and B.A prepared the manuscript. E.M.C, F.B.K., F.L-G, S.T-S, R.L-P, B.M, M.R-M, D.C-A., A.L-G, S.G-M., M.A.M., M.P-M., E.R-O., F.S-S., J.C-R., JF.Q-E., MT.S-C., J.S-P., R.B-F., M.I-G., I.R-A., MI.A., R.B-L., B.A-B., J.E., JJ.G-P., B.G-F., C.G-L., N.I., V.L., I.M., I.M., B.M-M., MS.R-L., M.G-H., P.P-M., J.L-P., S.A-A., S.A.dA., A.F-J., I.L-R., B.G., C.A., A.A-L., M.P-B., A.C., I.V., AF.M-M., I.L-S contributed to the acquisition, analysis and interpretation of the data used in the study. All authors critically reviewed the present manuscript, approved this final version and are accountable for all aspects of the work.

