## Supplementary Material for "Clinical description, molecular delineation and genotype-phenotype correlation in 340 patients with KBG syndrome: Addition of 67 new patients"

**1. Phenotyping questionnaire**

1. Demographic data

- Gender (Male, Female)
- Age (years) (by 2021)
- Ethnicity

2. Personal history

- Type of conception (Spontaneous/assisted)
- Intrauterine growth restriction (yes/no)
- Birth length and percentile
- Birth weight and percentile
- Pregnancy and perinatal complications (describe)

3. Medical investigations

- Magneticresonanceimaging (MRI) (normal/abnormal, describe)
- Electroencephalography (EEG) (normal/abnormal, describe)
- Blood tests (normal/abnormal, describe)
- Delayed bone age (yes/no)

4. Neurodevelopment

- Age at independent walking (months)
- Age at first words (months)
- Motor delay (yes/no)
- Language delay (yes/no)
- Intellectual dissability (yes/no)
- Autism spectrum disorder (ASD) diagnosis or autistic features (yes/no)
- Attention deficit hyperactivity disorder (ADHD) diagnosis or features (yes/no)

**5. Neurologicfindings**

- Sleep disorders (yes/no)
- Hypotonia (yes/no)
- Seizures (yes/no and type)
- Delayed closure or enlargement of fontanelle (yes/no)

**6. Otherclinicalfindings**

- Cardiopathy (yes/no and type)
- Hearing loss (yes/no and type)
- Recurrent otitis media
- Visual impairment (yes/no and type)
- Feeding difficulties (yes/no)
- Cryptorchidism (yes/no)

7. Physicalexamination

- Stature and percentile
- Cephalic perimeter and percentile
- Hand anomalies (yes/no and type)
- Costovertebral anomalies (yes/no and type)
- Head shape anomalies (yes/no and type)
- Low hairline (yes/no)
- Short or webbed neck (yes/no)
- Triangular face (yes/no)
- Microretrognathia (yes/no)
- Prognatism (yes/no)
- Macrostomy (yes/no)
- Lips
  - Shape: thin, thick/lower/upper
  - Cupid’sbow (yes/no)
- Macrodontia (yes/no) and or dental anomalies (yes/no and type)
- Philtrum (long/short, prominent/smooth)
- Strabismus (yes/no)
- Eyebrows (wide/bushy synophrys, yes/no)
- Hypertelorism (yes/no)
- Palpebral fissures (describe)
- Anteverted nares (yes/no)
- Bulbous nasal tip (yes/no)
- Prominent nose (yes/no)
- Ears:
  - Large or prominent
  - Low-set
- Other phenotypic feature (describe)

8. Genetic investigations

- Genetics tests performed (describe) and technologies used
- Diagnosis of suspicion (describe)
- ANKRD11 variant
- Paternal Origin: De novo variant (yes/no)

**2. Participating centers and number of patients recruited per center**

| **Center** | **N** |
| --- | --- |
| Fundación Jimenéz Díaz University Hospital | 12 |
| Sant Joan de DeuUniversity Hospital | 8 |
| Valld'HebronUniversity Hospital | 8 |
| La Paz University Hospital | 8 |
| 12 de OctubreUniversity Hospital | 7 |
| Cruces University Hospital | 5 |
| Quirón Salud Madrid University Hospital | 4 |
| Navarra UniversityClinic | 3 |
| Salamanca University Hospital | 3 |
| Miguel Servet University Hospital | 2 |
| Virgen de la ArrixacaUniversity Hospital | 1 |
| Asturias Central University Hospital | 1 |
| Getafe University Hospital | 1 |
| Santiago University Hospital | 1 |
| Niño Jesús University Hospital | 1 |
| Son LlátzerUniversity Hospital | 1 |
| Can MissesUniversity Hospital | 1 |
| Barcelona Clínic Hospital | 1 |

**3. Studies from the literature review and number of patients included**

| **Paper (first author and year)** | **PMID** | **N** |
| --- | --- | --- |
| Kutkowska-Kazmierczak et al, 2021 | 34440431 | 23 |
| Mattei et al, 2021 | 33494799 | 1 |
| Parenti et al, 2021 | 33955014 | 23 |
| Nardello et al, 2021 | 33476899 | 1 |
| Bucerzan et al, 2020 | 32760686 | 1 |
| Cucco et al, 2020 (Patient B) | 32476269 | 1 |
| Gnazzo et al, 2020 | 32124548 | 31 |
| Jin Kim et al, 2020 | 33262785 | 2 |
| Reuter et al, 2020 | 32037394 | 1 |
| Sayed et al, 2020 | 32222090 | 2 |
| Alves et al, 2019 | 30642272 | 1 |
| Libianto et al, 2019 | 30877071 | 1 |
| VanDongen et al, 2019 | 30786142 | 12 |
| Behnert et al, 2018 | 29696793 | 1 |
| Bianchi et al, 2018 | 29224748 | 1 |
| DeBernardi et al, 2018 | 30088855 | 1 |
| Low et al, 2017 | 28815928 | 1 |
| Miyatake et al, 2017 | 28250421 | 3 |
| Murray et al, 2017 | 28449295 | 14 |
| Novara et al, 2017 | 28422132 | 11 |
| Srivastava et al, 2017 | 28099180 | 1 |
| Goldenberg et al, 2016 | 27605097 | 38 |
| Kleyner et al, 2016 | 27900361 | 1 |
| Low et al, 2016 | 27667800 | 33 |
| Palumbo et al, 2016 | 32604767 | 1 |
| Parenti et al, 2016 | 25652421 | 2 |
| Crippa et al, 2015 | 25838844 | 3 |
| Kim et al, 2015 | 25464108 | 3 |
| Ockeloen et al, 2015 | 26269249 | 20 |
| Walz et al, 2015 | 25413698 | 6 |
| Lim et al, 2014 | 25187894 | 1 |
| Khalifa et al, 2013 | 23494856 | 2 |
| Miyatake et al, 2013 | 23463723 | 1 |
| Scarano et al, 2013 | 31191201 | 12 |
| Spengler et al, 2013 | 23885231 | 1 |
| Isrie et al, 2012 | 21654729 | 2 |
| Sacharow et al, 2012 | 22307766 | 2 |
| Sirmaci et al, 2011 | 21782149 | 7 |
| Youngs et al, 2011 | 21527850 | 1 |
| Willemsen et al, 2010 | 19920853 | 4 |
